# Interest in and feasibility of a dementia prevention program among community-dwelling older adults: a questionnaire survey

**DOI:** 10.64898/2026.03.22.26349026

**Authors:** Minoru Kouzuki, Hideyuki Tazumi, Nao Nakada

## Abstract

**Background:** Evidence regarding dementia prevention strategies has been accumulating. However, disseminating research findings to the public is often difficult, and addressing the evidence–practice gap presents an important challenge. This study examined potential strategies to support sustained engagement in dementia prevention activities.

**Participants and Setting:** Members of senior citizens’ clubs in Tottori Prefecture, Japan.

**Methods:** This questionnaire survey collected data on basic demographics, frailty, and subjective cognitive decline (SCD). It also included questions on awareness of the Tottori Method Dementia Prevention Program, interest in experiencing the program if an instructor was dispatched, and the feasibility of engaging in the program through internet-based delivery or printed materials.

**Results:** A total of 9,506 respondents were included in the analysis. Awareness of the dementia prevention program was 11.9%. Overall, 50.4% of the respondents registered a desire to try the program if an instructor was dispatched. The highest proportion of respondents (50.5%) reported willingness to engage in the program if materials summarizing activities that could be completed in approximately 10 min were provided. However, both frailty and SCD were associated with a lower interest in these dementia prevention activities (adjusted odd ratio [95% confidence interval] = 0.77 [0.67–0.89] and 0.86 [0.79–0.95], respectively).

**Conclusions:** To promote sustained engagement in dementia prevention activities, opportunities to experience the program and activities that can be completed in a short time should be availed. However, disseminating research findings to the public remains challenging, and individuals at a higher risk of health problems may be less interested in dementia prevention. Proactive outreach strategies targeting high-risk individuals may be necessary to effectively disseminate the information.

**KEY POINTS BOX:** *Key points:*

- Disseminating research findings to the public is challenging.
- To sustain dementia prevention activities, individuals should learn through hands-on experience and the activities should take a short time to complete.
- Individuals at higher risk of health problems, such as those with frailty or subjective cognitive decline, tend to have lower interest in dementia prevention.

*Why does this paper matter?:* This study identified practical approaches for encouraging sustained engagement in dementia prevention activities and highlighted challenges in providing information to the public. The findings provide important insights into designing dementia prevention research programs and disseminating research outcomes to guide social implementation of dementia prevention strategies.

## INTRODUCTION

Multifactorial interventions focusing on lifestyle factors have been widely recognized as effective at preventing cognitive decline, and intervention studies have consequently been conducted worldwide.^1^ In 2017, the lead author, together with the research collaborators, developed a program to prevent cognitive decline incorporating physical exercise, cognitive training, and educational lectures on dementia and lifestyle habits (named the “Tottori Method Dementia Prevention (TMDP) Program”).^2^ The program comprises weekly 2-h sessions (50 min of physical exercise, 20 min of lectures or breaks, and 50 min of cognitive training) conducted for 6 months among community-dwelling adults aged ≥65 years who were suspected of having mild cognitive decline. Participation in the program improved both cognitive and physical functions.^2^

The TMDP Program has been disseminated since fiscal year 2019 through public lectures, television, newspapers, and prefectural websites. However, translating research findings into real-world practice is often challenging. Even when an intervention has demonstrated efficacy under ideal conditions in a research setting, various barriers can arise during implementation in real-world contexts,^3^ resulting in a gap between evidence and practice.

Community residents require a support system to sustain dementia prevention activities. Owing to limited human resources, more efficient support strategies need to be explored. In recent years, digital tools have attracted attention as potential means of providing effective support under limited social resources. Digital skills may act as health determinants among older adults through social compensation, health intelligence, and technological empowerment,^4^ suggesting that digital tools may not only serve as a support method but also contribute to improvements in health behaviors. Although the digital divide among older adults has been decreasing, it still persists.^5^ In addition, older adults show relatively low positive attitudes to health apps,^6^ and some individuals who wish to improve their health are not interested in using digital tools.^7^ Therefore, careful consideration is necessary when evaluating whether digital tools represent a realistic approach for supporting today’s older population.

In disseminating dementia prevention activities, the choice of delivery method is an important issue. However, few studies have examined the demand for different delivery methods of dementia prevention activities and analyzed these preferences according to participant characteristics. Consequently, the most effective and efficient approaches for supporting dementia prevention activities remain unclear.

Therefore, in the present study, we investigated the demand for different methods of delivering dementia prevention activities using a questionnaire survey and analyzed their associations with participant characteristics. We further examined the potential strategies for supporting sustained engagement in dementia prevention activities.

## METHODS

### Study Design

This study followed a cross-sectional design and was guided by the Strengthening the Reporting of Observational Studies in Epidemiology (STROBE) checklist (Supplementary Appendix).^8^

### Participants and data collection

A questionnaire survey was conducted between September and October 2024 among members of senior citizens’ clubs in Tottori Prefecture, Japan. As of April 2024, 570 senior citizens’ clubs existed in Tottori Prefecture with a total membership of 23,651 individuals.^9^ Tottori Prefecture had a population of approximately 530,000 as of April 2024^10^ and is the least populous prefecture in Japan. Tottori Prefecture comprises 19 municipalities, with a large variation in population size; the most populous municipality has approximately 183,000 residents, while the least populous has approximately 2,400 residents.^10^ Members of senior citizens’ clubs are present in all municipalities; however, municipalities with larger populations tend to have more members, while those with smaller populations tend to have fewer members.

The procedures for the questionnaire survey were as follows:

1. Research request letters and questionnaire forms corresponding to the number of members in each unit club were sent to the respective presidents of the individual senior citizens’ clubs in Tottori Prefecture.
2. Each club president distributed the research request letters and questionnaire forms to the members.
3. Club members who consented to participate in the research submitted completed questionnaires to their respective club presidents.
4. The club presidents then mailed the completed questionnaires to the research team (Tottori Prefecture Federation of Senior Citizens Clubs).

A total of 10,063 questionnaires were returned. Among them, 557 questionnaires without a check in the consent confirmation box were excluded. Consequently, 9,506 questionnaires were included in the analysis. When responses to specific items were incomplete, those items were treated as missing values, and only the available data were used in the analyses.

### Ethics and informed consent

This study was approved by the Ethical review board, Faculty of Medicine Tottori University (No. 24A074). Participants were informed about the study using a research request letter, and consent was obtained by checking the consent confirmation box on the questionnaire response form.

### Questionnaire survey

The questionnaire comprised the following components (Supplementary Table S1):

1. demographic data, such as age and sex;
2. assessment of frailty;
3. assessment of subjective cognitive decline (SCD);
4. engagement in dementia prevention activities;
5. internet environment at home;
6. use of online tools, such as web conferencing, LINE, or YouTube;
7. awareness of the TMDP Program, interest in experiencing the program if an instructor was dispatched, and the feasibility of participating through internet-based delivery or printed materials;
8. questions regarding the participant’s residential area.

Participants were asked to describe their residential area in free text, whereas the other items were answered by selecting from predefined response options. To allow respondents unfamiliar with the TMDP Program to answer the questionnaire, a brief description of the program and its previously reported outcomes was included in the questionnaire (Supplementary Table S1).

Frailty and SCD were assessed based on the questionnaire responses. Frailty status was determined using five questions designed to assess frailty (nutrition/shrinking in Q3, physical function in Q4, physical activity in Q5, emotions/exhaustion in Q6, and forgetfulness in Q7), following previous studies.^11^ One point was assigned for a “yes” response to the questions on nutrition/shrinking, physical function, and emotions/exhaustion, and one point was assigned for a “no” response to the questions on physical activity and forgetfulness. The total score was calculated and categorized as frailty (≥3 points), prefrailty (1–2 points), or robust (0 points). SCD was also assessed based on previous studies.^12^ Participants were classified as having SCD if they answered “yes” to either the forgetfulness question (Q8) or time-orientation question (Q9).

### Statistical analysis

Logistic regression analyses were conducted to examine the associations between questionnaire results on the TMDP Program and participant characteristics. Dependent variables were derived from questions related to the TMDP Program. Independent variables included age, sex, frailty status, SCD status, engagement in dementia prevention activities, internet environment at home, and experience using online tools. For each model, one of these variables was selected as the main explanatory variable. Furthermore, a multiple logistic regression analysis was conducted, with variables considered as related to the dependent variables additionally included as covariates. Finally, odds ratios (ORs) or adjusted ORs (aORs) and their 95% confidence intervals (CIs) were calculated. Given the low frequency of missing values (<10%), sensitivity analyses were not considered necessary for this cross-sectional analysis. Statistical analyses were performed using EZR statistical software (version 1.55, Saitama Medical Center, Jichi Medical University, Saitama, Japan),^13^ and statistical significance was set at p < 0.05.

## RESULTS

### Participant characteristics

Participant characteristics are shown in Table 1. Individuals aged ≥65 years accounted for 95.2% of the sample. Regarding frailty status, prefrail individuals constituted the largest group. When prefrail and frail individuals were combined, they accounted for 81.4% of the total sample. The prevalence of SCD was 33.1%, 21.4% of respondents engaged in dementia prevention activities. The proportion of participants with internet access at home was 51.8%; further, with respect to online communication, 10.0% had experience using web conferencing systems, 46.5% using LINE, and 25.7% using YouTube.

**Table 1.**
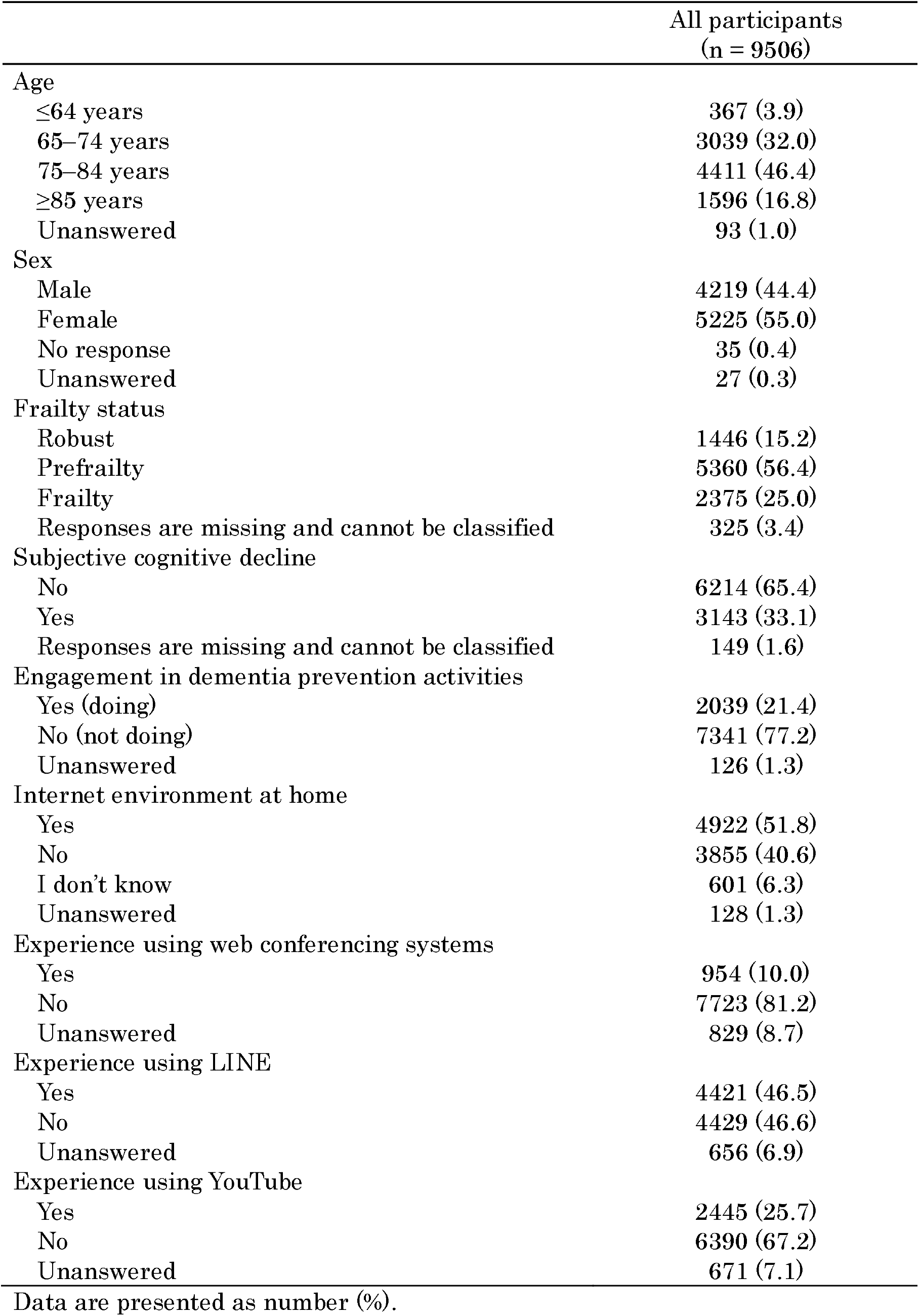
Characteristics of the participants.

|  | All participants<br>(n = 9506) |
| --- | --- |
| Age |  |
| ≤64 years | 367 (3.9) |
| 65–74 years | 3039 (32.0) |
| 75–84 years | 4411 (46.4) |
| ≥85 years | 1596 (16.8) |
| Unanswered | 93 (1.0) |
| Sex |  |
| Male | 4219 (44.4) |
| Female | 5225 (55.0) |
| No response | 35 (0.4) |
| Unanswered | 27 (0.3) |
| Frailty status |  |
| Robust | 1446 (15.2) |
| Prefrailty | 5360 (56.4) |
| Frailty | 2375 (25.0) |
| Responses are missing and cannot be classified | 325 (3.4) |
| Subjective cognitive decline |  |
| No | 6214 (65.4) |
| Yes | 3143 (33.1) |
| Responses are missing and cannot be classified | 149 (1.6) |
| Engagement in dementia prevention activities |  |
| Yes (doing) | 2039 (21.4) |
| No (not doing) | 7341 (77.2) |
| Unanswered | 126 (1.3) |
| Internet environment at home |  |
| Yes | 4922 (51.8) |
| No | 3855 (40.6) |
| I don't know | 601 (6.3) |
| Unanswered | 128 (1.3) |
| Experience using web conferencing systems |  |
| Yes | 954 (10.0) |
| No | 7723 (81.2) |
| Unanswered | 829 (8.7) |
| Experience using LINE |  |
| Yes | 4421 (46.5) |
| No | 4429 (46.6) |
| Unanswered | 656 (6.9) |
| Experience using YouTube |  |
| Yes | 2445 (25.7) |
| No | 6390 (67.2) |
| Unanswered | 671 (7.1) |

Data are presented as number (%).

### Results related to the TMDP program

The results related to the TMDP Program are shown in Figure 1. The proportion of participants aware of the TMDP Program was low at 11.9%. When analyzed by residential area, the median rate was 13.6% (minimum 3.3%, maximum 25.4%). Among the participants, 50.4% expressed the desire to experience the program at a club gathering if an instructor was dispatched. When analyzed by residential area, the median rate was 51.7% (minimum 38.6%, maximum 62.9%). If the program was delivered online, 36.4% and 36.3% reported willingness to engage in the program at a club gathering and at home, respectively, showing no substantial difference. By residential area, the median proportions were 37.0% (minimum 21.7%, maximum 52.4%) and 37.7% (minimum 25.3%, maximum 47.9%), respectively. If printed materials were provided, 46.1% reported willingness to regularly engage in the program at club gatherings. When the program was described as taking approximately 10 min per session, the proportion increased slightly to 50.5%. By residential area, the median proportions were 46.3% (minimum 39.8%, maximum 66.7%) and 51.5% (minimum 44.6%, maximum 70.0%), respectively.

**Figure 1.**
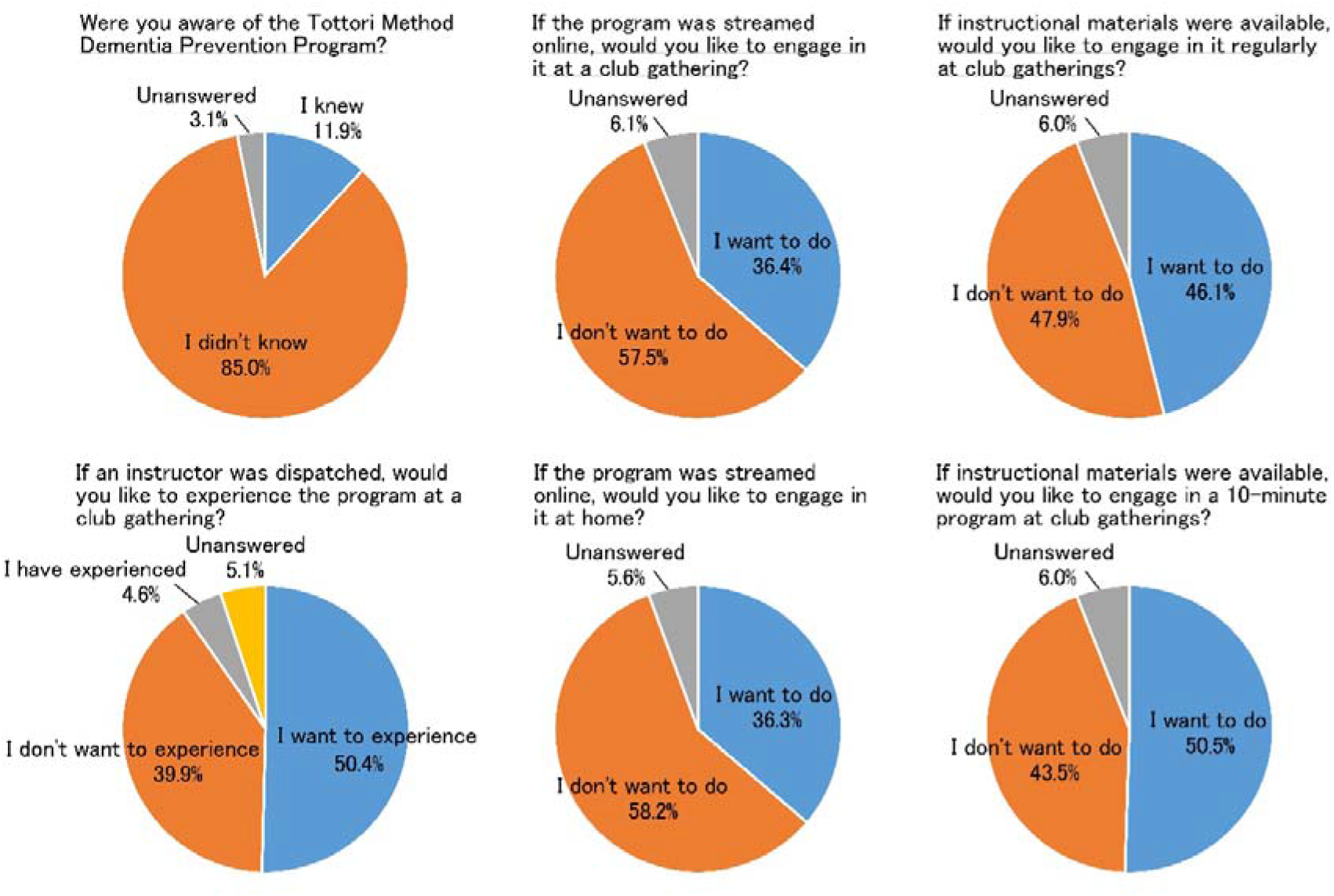
Questionnaire survey results on the dementia prevention program.

Next, factors associated with responses related to the TMDP Program were examined (Table 2, Supplementary Tables S2–S10). Frailty was significantly associated with unawareness of the TMDP Program (aOR = 0.80, 95% CI = 0.64–0.99) and unwillingness to experience the program even if an instructor was dispatched (aOR = 0.74, 95% CI = 0.64–0.86), engage in club gatherings even if the program was delivered online (aOR = 0.81, 95% CI = 0.69–0.94), and engage in the program either regularly or for the 10 min shortened version, even if printed materials were provided (aOR = 0.71, 95% CI = 0.62–0.82; aOR = 0.77, 95% CI = 0.67–0.89). SCD was significantly associated with unawareness of the TMDP Program (aOR = 0.84, 95% CI = 0.72–0.98) and unwillingness to engage in the program either regularly or for the 10 min shortened version, even if printed materials were provided (aOR = 0.85, 95% CI = 0.77–0.93; aOR = 0.86, 95% CI = 0.79–0.95). Non-engagement in dementia prevention activities was significantly associated with unawareness of the TMDP Program (aOR = 0.32, 95% CI = 0.28–0.38) and unwillingness to experience the program even if an instructor was dispatched (aOR = 0.45, 95% CI = 0.40–0.51), engage even if the program was delivered online at club gatherings or at home (aOR = 0.55, 95% CI = 0.49–0.62; aOR = 0.58, 95% CI = 0.51–0.65), and engage in the program either regularly or for the 10 min shortened version, even if printed materials were provided (aOR = 0.51, 95% CI = 0.46–0.57; aOR = 0.50, 95% CI = 0.45–0.56).

**Table 2.**
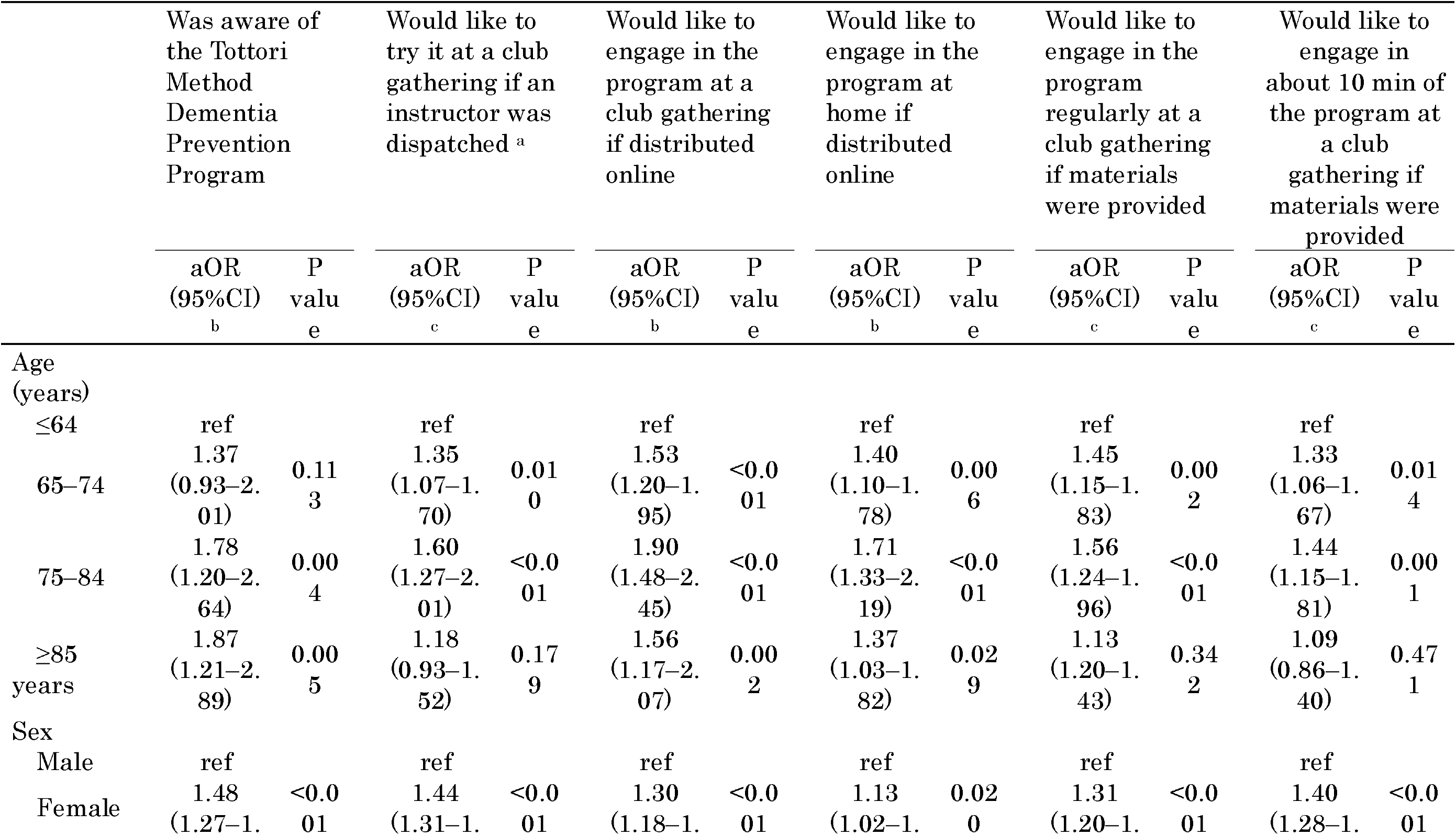
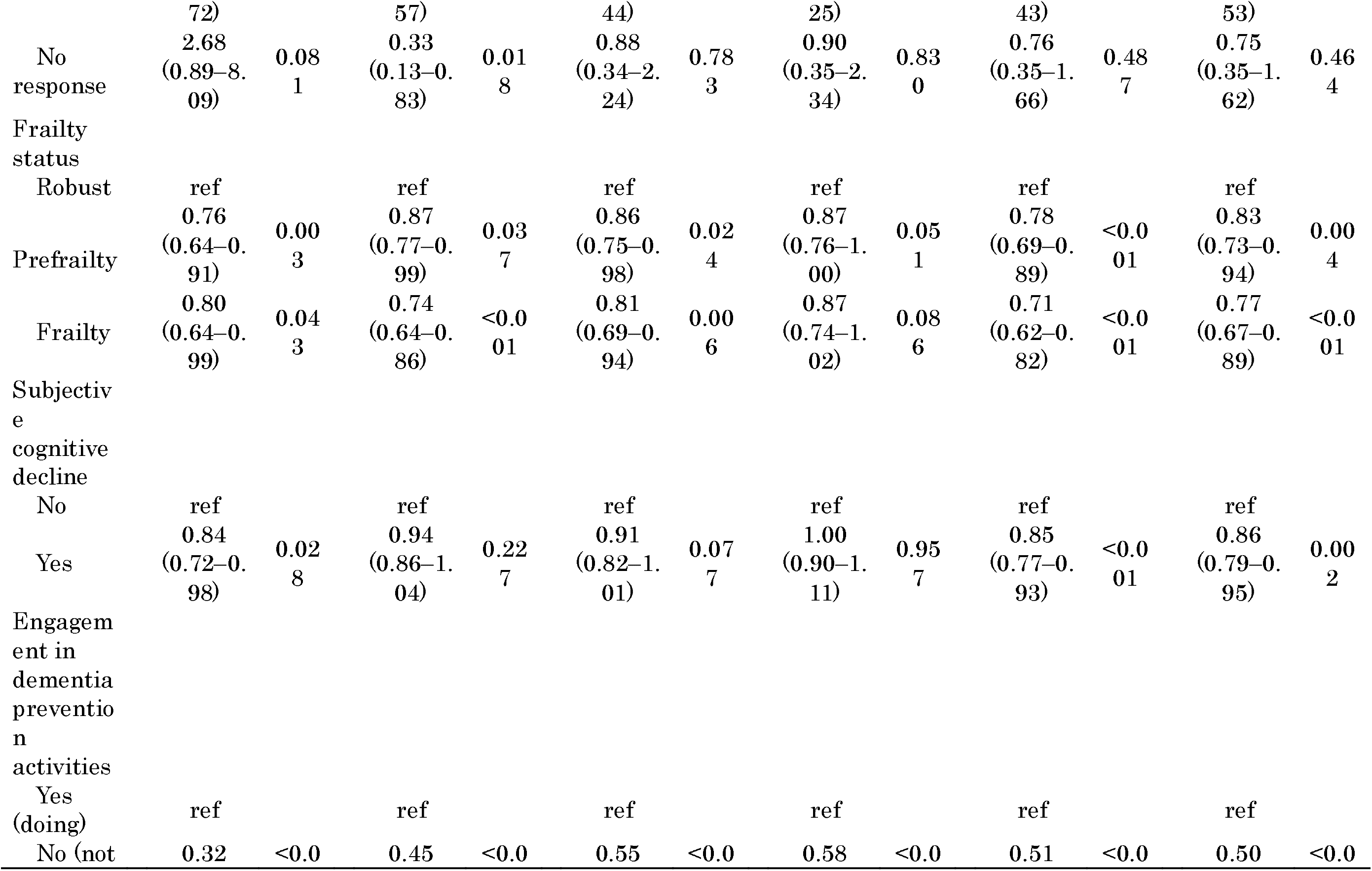
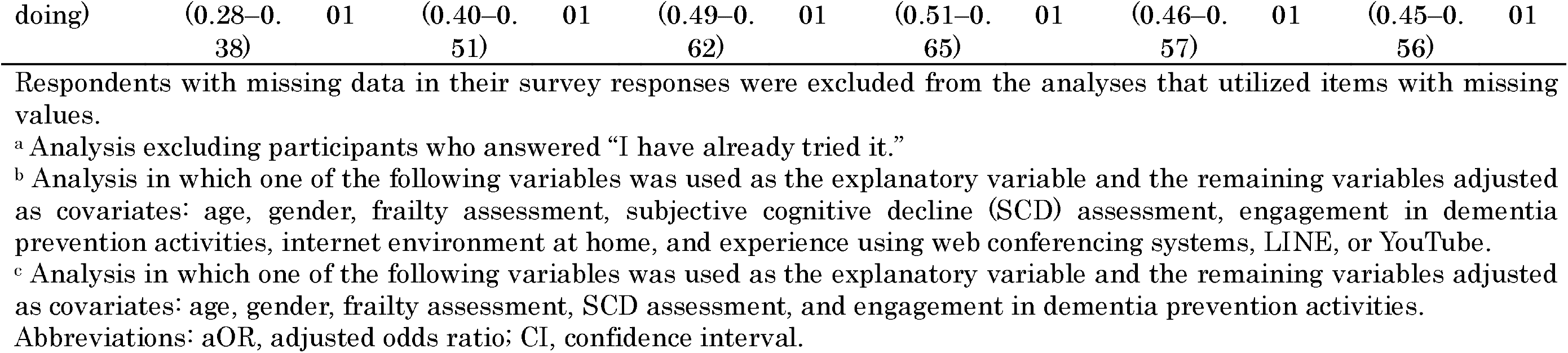
Association between results of the questionnaire on the dementia prevention program and participant characteristics multiple logistic regression analyses)

## DISCUSSION

To our knowledge, this study is the first to examine demand for different delivery methods of dementia prevention activities according to participant characteristics and provide insights into the implementation of research output in real-world settings. A questionnaire survey regarding the implementation of the TMDP Program—a cognitive decline prevention program—was conducted among community-dwelling residents. More than half of the respondents reported a desire to experience the program if an instructor was dispatched and showed willingness to engage in the program if materials summarizing the activities that could be completed in approximately 10 min were available. These findings suggest that providing opportunities for hands-on experience and offering activities that can be performed in a short time are key factors for promoting sustained engagement in dementia prevention activities. However, disseminating research findings to the public remains challenging, and individuals at higher risk of health problems may be less interested in dementia prevention activities. Therefore, in addition to continuous population-based approaches, proactive outreach strategies targeting high-risk individuals may be necessary.

First, the difficulty of disseminating research findings was clearly demonstrated. Only 11.9% of respondents were aware of the TMDP Program despite dissemination efforts over more than 5 years, indicating a relatively low level of awareness. The awareness rates also varied by region, ranging from 3.3% to 25.4%. Rural residents are reported to have lower access to health information from sources such as primary care providers, specialist doctors, blogs, and magazines compared with residents in urban areas.^14^ Because county-level disadvantage has been associated with the risk of developing frailty and probable dementia,^15^ dissemination efforts should consider geographic and social contexts.

Half of the respondents (50.4%) expressed the desire to experience the program at senior citizens’ club gatherings if an instructor was dispatched, suggesting a certain level of demand. A previous systematic review on physical activity programs reported an initial participation rate of 9.2% and sustained participation rate of 79.8%^16^. Therefore, providing opportunities for individuals to first experience the program may be particularly meaningful. However, sustained practice requires autonomy and habit formation. When asked about online delivery, approximately 36% of the respondents expressed willingness to engage in the program either at club gatherings or at home, suggesting no clear difference in demand between group-based and individual participation. Notably, the demand for such approaches was influenced by the availability of internet at home and prior experience using online tools (Supplementary Table S9, 10), indicating that digital-based interventions may be unsuitable for individuals unfamiliar with digital technologies. Furthermore, some individuals report concerns about operating digital tools for health promotion,^7^ suggesting the need for learning opportunities on the use of online tools to facilitate wider adoption of digital approaches.

In contrast, the distribution of printed materials does not require explanations on usage. When asked whether they would be willing to engage in approximately 10 min of physical exercise or cognitive training during club gatherings if materials were available, 50.5% of respondents answered affirmatively, representing the highest demand among the support methods examined in this study. In a previous study involving adults aged ≥60 years with cognitive impairment, an intervention based the nudge theory that delivered content through posters, information brochures, and websites significantly improved cognitive health behaviors and cognitive performance.^17^ Thus, distributing printed materials may promote awareness of dementia prevention behaviors to a large target population without the need to engage specialized professionals. In addition, engagement in leisure activities involving physical, cognitive, and social activities has been associated with a reduced risk of dementia.^18^ Of note, regardless of the effectiveness of a program, participation must be sustained to accrue benefits. Because exercise detraining has been shown to negatively affect cognitive function,^19^ strategies that promote habit formation through brief but continuous activities may be effective. Multidomain lifestyle interventions have also demonstrated improvements in lifestyle risk factors related to Alzheimer’s disease and cognitive function,^20^ and midlife-initiated physical activity has been associated with reduced risk of dementia and cognitive impairment in later life.^21^ These findings underscore the importance of embracing healthy lifestyle behaviors at an early age.

Individuals not currently engaged in dementia prevention activities, those suspected of frailty, and those suspected of having SCD—groups at a higher risk of health problems—tended to show lower interest in dementia prevention activities. Although a population approach is necessary to widely disseminate dementia prevention initiatives, those who most need to engage in preventive activities may not show interest in them. In a 6-year dementia prevention intervention study, older age, poorer cognition, more symptoms of depression, and greater disability were associated with dropout from the trial.^22^ Therefore, in addition to broadly disseminating information, the health status of high-risk individuals should be assessed for provision of targeted support. Furthermore, low health literacy among older adults was negatively associated with health behaviors,^23^ underscoring the need to improve health literacy. The Health Belief Model, a widely used theory of health behavior, proposes that behaviors are influenced by perceived susceptibility, perceived severity, perceived benefits, perceived barriers, self-efficacy, and cues to action.^24^ A previous study reported that cues to action and self-efficacy influence dementia preventive behaviors.^25^ Moreover, multidomain lifestyle interventions considered beneficial for cognition^26^ have also shown effectiveness in preventing or reversing frailty.^27,28^ Because social prejudice regarding dementia still exists,^29^ it is important to provide accurate information, eliminate misconceptions, and promote behavioral change by emphasizing not only the benefits of dementia prevention but also other health benefits and feasibility of the activities.

Finally, some limitations should be acknowledged. First, since the methods used to disseminate the research findings related to the TMDP Program were not strictly regulated, the specific dissemination strategies underlying the observed results cannot be easily determined. Nevertheless, considering that the program has been introduced in various settings since fiscal year 2019, the findings illustrate the difficulty of disseminating research outcomes to the public. Second, this study was based on a questionnaire survey. Because the dementia prevention program was not actually delivered in this study, it remains unclear whether the participants would continue the activities if provided using their preferred methods. Third, the results may differ according to regional characteristics. Tottori Prefecture has the smallest population in Japan and a relatively high aging rate (approximately 34% in 2024). Because online activities are highest in metropolitan residents compared with rural residents,^30^ surveys conducted in urban settings may yield a higher demand for interventions delivered through online platforms.

In conclusion, this study demonstrated that informing the general public about research outcomes related to dementia prevention is a challenging endeavor. As potential support strategies for promoting sustained engagement in dementia prevention activities, notable demand was observed for opportunities to experience the program and materials describing shortened activities. However, individuals with a higher risk of health problems, such as those with frailty or SCD, had lower interest in dementia prevention activities. Therefore, in addition to broad information dissemination, proactive outreach strategies tailored to participant characteristics should be provided. Future studies should examine whether information dissemination leads to sustained engagement in dementia prevention activities and improvements in health outcomes.

## Data Availability

The datasets used and/or analyzed during the current study are available from the corresponding author upon reasonable request, subject to the approval of all authors, relevant parties, and the ethics committee.

## ACKNOWLEDGMENTS

We would like to thank all the individuals who participated in this study and express gratitude to the staff of the Tottori Prefecture Federation of Senior Citizens Clubs. We also thank Editage (www.editage.com) for the English language editing.

## Author Contributions

Study concept and design: All authors. Acquisition of data: Minoru Kouzuki. Analysis of data: Minoru Kouzuki. Interpretation of data: All authors. Draft of the manuscript: Minoru Kouzuki. Reviewed the manuscript: All authors. Final approval of the published version: All authors.

## Declaration of generative AI and AI-assisted technologies in the writing process

ChatGPT (OpenAI) and Gemini (Google) were used to assist in the drafting of this manuscript. The authors critically reviewed and revised the output and take full responsibility for the final content.

## Supplemental Material

Additional supporting information can be found online in the Supporting Information section.

**Supplementary Table S1:** English translation of the relevant questions used in this study.

**Supplementary Table S2:** Responses to the question “Did you know about the Tottori Method Dementia Prevention Program?” according to participant characteristics.

**Supplementary Table S3:** Responses to the question “If an instructor was dispatched, would you like to try the program at a club gathering?” according to participant characteristics.

**Supplementary Table S4:** Responses to the question “If the program was distributed online, would you like to engage in it at a club gathering?” according to participant characteristics.

**Supplementary Table S5:** Responses to the question “If the program was distributed online, would you like to engage in it at home?” according to participant characteristics.

**Supplementary Table S6:** Responses to the question “If materials describing how to perform the program were provided, would you like to engage in it regularly at a club gathering?” according to participant characteristics.

**Supplementary Table S7:** Responses to the question “If materials describing how to perform the program were provided, would you like to engage in a program of about 10 min at each club gathering?” according to participant characteristics.

**Supplementary Table S8:** Association between results of the questionnaire on the dementia prevention program and participant characteristics (univariate logistic regression analysis).

**Supplementary Table S9:** Association between results of the questionnaire on the dementia prevention program and home internet environment and the use of online tools (univariate logistic regression analysis).

**Supplementary Table S10:** Association between results of the questionnaire on the dementia prevention program and home internet environment and the use of online tools (multiple logistic regression analysis).

**Supplementary Appendix:** STROBE Statement—Checklist of items that should be included in reports of cross-sectional studies.

