## Supplemental Material for "Interest in and feasibility of a dementia prevention program among community-dwelling older adults: a questionnaire survey"

Table S1. English translation of the relevant questions used in this study
(The questions are translated from Japanese)

Please circle the option that best applies to each of the following questions. Some questions may be difficult to answer, but if you are unsure, please select the option that best fits “If I was to say it.”

Q1. Please tell us your age.
→ ① ≤64 years・ ② 65–74 years ・ ③ 75–84 years ・ ④ ≥85 years

Q2. Please tell us your sex.
→ ① Male ・ ② Female ・ ③ No response

Q3. Have you lost 2–3 kg in the past 6 months?
→ ① Yes ・ ② No

Q4. Do you think you walk slower than before?
→ ① Yes ・ ② No

Q5. Do you go for a walk for your health at least once a week?
→ ① Yes ・ ② No

Q6. In the past 2 weeks, have you felt tired without a reason?
→ ① Yes ・ ② No

Q7. Can you recall what happened 5 min ago?
→ ① Yes ・ ② No

Q8. Do your family or your friends point out your memory loss (e.g., "You're always asking the same thing")?
→ ① Yes ・ ② No

Q9. Do you find yourself not knowing today's date?
→ ① Yes ・ ② No

Q10. Are you currently taking any actions to prevent dementia?
→ ① Yes ・ ② No

Q11. Do you have internet environment at home (wireless LAN such as Wi-Fi)?
→ ① Yes ・ ② No ・ ③ I don’t know

Q12. Please indicate which of the following you have used for online communication.

1. Experience using a web conferencing system (e.g., Zoom)
   → ① Yes ・ ② No
2. Experience using LINE
   → ① Yes ・ ② No
3. Experience using YouTube
   → ① Yes ・ ② No

The Tottori Prefecture Federation of Senior Citizens Clubs is working to promote and raise awareness of the Tottori Method Dementia Prevention Program, which was developed in Tottori Prefecture. This program comprises physical exercise for dementia prevention (such as aerobic and strength training), cognitive training (such as memorization and calculation tasks that stimulate the brain), and educational lectures (classes about dementia). The program lasts 2 hours and is used in community-based preventive care classes and similar settings. In many cases, specialists such as occupational therapists provide instructions when the program is implemented; however, the program is designed with simple content for ease of teaching by non-specialists. To evaluate its efficacy, individuals with mild cognitive decline participated in the program once a week for 6 months. The results showed improvements in cognitive function and physical function.

Q13. Were you aware of the "Tottori Method Dementia Prevention Program"?
→ ① Yes, I knew about it ・ ② No, I did not know about it

Q14. If a specialist who can provide instructions on the "Tottori Method Dementia Prevention Program" was dispatched, would you like to try it during a gathering of your senior citizens' club? Please note that the program duration can be shortened to about one hour to match the gathering time; therefore, please answer without worrying about the time required.
→ ① I would like to try it ・ ② I would not like to try it ・ ③ I have already tried it

Q15. If the "Tottori Method Dementia Prevention Program" was distributed via the internet (e.g., using Zoom, YouTube, or LINE), would you like to engage in it during a senior citizens' club gathering? Please answer based on your own feelings without considering whether the gathering venue has internet access or whether someone is familiar with operating devices such as computers or smartphones.
→ ① I would like to engage ・ ② I would not like to engage

Q16. If the "Tottori Method Dementia Prevention Program" was distributed via the internet (e.g., using Zoom, YouTube, or LINE), would you like to engage in it at home? Please answer based on your own feelings without considering whether your home has internet environment or whether you can operate devices such as computers or smartphones.
→ ① I would like to engage ・ ② I would not like to engage

Q17. If materials such as a guide explaining how to perform the program activities were distributed, would you like to regularly engage in the exercise and cognitive activities of the "Tottori Method Dementia Prevention Program" by yourself during a senior citizens' club gathering?
→ ① I would like to engage ・ ② I would not like to engage

Q18. If materials such as a guide explaining how to perform the program activities were distributed, would you like to engage in about 10 min of the exercise and cognitive activities of the "Tottori Method Dementia Prevention Program" at each senior citizens' club gathering by yourself?
→ ① I would like to engage ・ ② I would not like to engage

Finally, please tell us the municipality where you live.
→ ________________ City・Town・Village
(For example, please write the municipality name such as "Yonago City.")

Table S2: Responses to the question “Did you know about the Tottori Method Dementia Prevention Program?” according to participant characteristics

| | Did you know about the Tottori Method Dementia Prevention Program? | |
|---|---|---|
| | Yes | No |
| Age | | |
| ≤64 years | 38 (3.4) | 324 (4.0) |
| 65–74 years | 336 (30.0) | 2652 (33.1) |
| 75–84 years | 573 (51.2) | 3698 (46.1) |
| ≥85 years | 172 (15.4) | 1344 (16.8) |
| Sex | | |
| Male | 405 (35.7) | 3724 (46.2) |
| Female | 724 (63.9) | 4310 (53.5) |
| No response | 4 (0.4) | 29 (0.4) |
| Frailty status | | |
| Robust | 233 (20.9) | 1185 (15.2) |
| Prefrailty | 621 (55.7) | 4590 (58.8) |
| Frailty | 260 (23.3) | 2037 (26.1) |
| Subjective cognitive decline | | |
| No | 800 (71.3) | 5244 (65.8) |
| Yes | 322 (28.7) | 2723 (34.2) |
| Engagement in dementia prevention activities | | |
| Yes (doing) | 485 (43.5) | 1482 (18.5) |
| No (not doing) | 629 (56.5) | 6508 (81.5) |
| Internet environment at home | | |
| Yes | 694 (61.5) | 4137 (51.8) |
| No | 393 (34.8) | 3316 (41.5) |
| I don’t know | 42 (3.7) | 530 (6.6) |
| Experience using web conferencing systems | | |
| Yes | 177 (17.2) | 764 (10.3) |
| No | 855 (82.8) | 6662 (89.7) |
| Experience using LINE | | |
| Yes | 641 (60.4) | 3707 (49.0) |
| No | 421 (39.6) | 3852 (51.0) |
| Experience using YouTube | | |
| Yes | 363 (34.2) | 2052 (27.2) |
| No | 697 (65.8) | 5500 (72.8) |

Data are presented as number (%).

Table S3: Responses to the question “If an instructor was dispatched, would you like to try the program at a club gathering?” according to participant characteristics

| | If an instructor was dispatched, would you like to try the program at a club gathering? | | |
|---|---|---|---|
| | Yes | No | I have already tried it |
| Age | | | |
| ≤64 years | 158 (3.3) | 195 (5.2) | 5 (1.1) |
| 65–74 years | 1513 (31.9) | 1313 (34.9) | 110 (25.3) |
| 75–84 years | 2340 (49.3) | 1600 (42.6) | 233 (53.6) |
| ≥85 years | 737 (15.5) | 651 (17.3) | 87 (20.0) |
| Sex | | | |
| Male | 1955 (40.9) | 1960 (51.8) | 155 (35.3) |
| Female | 2822 (59.0) | 1800 (47.6) | 280 (63.8) |
| No response | 6 (0.1) | 22 (0.6) | 4 (0.9) |
| Frailty status | | | |
| Robust | 796 (17.1) | 519 (14.1) | 86 (20.1) |
| Prefrailty | 2763 (59.4) | 2132 (57.8) | 236 (55.1) |
| Frailty | 1092 (23.5) | 1035 (28.1) | 106 (24.8) |
| Subjective cognitive decline | | | |
| No | 3182 (67.4) | 2462 (65.6) | 301 (69.8) |
| Yes | 1539 (32.6) | 1293 (34.4) | 130 (30.2) |
| Engagement in dementia prevention activities | | | |
| Yes (doing) | 1251 (26.5) | 493 (13.1) | 198 (45.7) |
| No (not doing) | 3465 (73.5) | 3276 (86.9) | 235 (54.3) |
| Internet environment at home | | | |
| Yes | 2715 (57.2) | 1829 (48.7) | 223 (51.5) |
| No | 1800 (37.9) | 1632 (43.5) | 179 (41.3) |
| I don’t know | 229 (4.8) | 293 (7.8) | 31 (7.2) |
| Experience using web conferencing systems | | | |
| Yes | 543 (12.3) | 347 (9.9) | 45 (11.2) |
| No | 3859 (87.7) | 3158 (90.1) | 355 (88.8) |
| Experience using LINE | | | |
| Yes | 2532 (56.2) | 1558 (43.8) | 212 (51.7) |
| No | 1976 (43.8) | 2002 (56.2) | 198 (48.3) |
| Experience using YouTube | | | |
| Yes | 1356 (30.2) | 934 (26.3) | 106 (25.9) |
| No | 3139 (69.8) | 2622 (73.7) | 303 (74.1) |

Data are presented as number (%).

Table S4: Responses to the question “If the program was distributed online, would you like to engage in it at a club gathering?” according to participant characteristics

| | If the program was distributed online, would you like to engage in it at a club gathering? | |
|---|---|---|
| | Yes | No |
| Age | | |
| ≤64 years | 135 (3.9) | 223 (4.1) |
| 65–74 years | 1205 (35.1) | 1726 (31.8) |
| 75–84 years | 1648 (48.1) | 2468 (45.5) |
| ≥85 years | 441 (12.9) | 1005 (18.5) |
| Sex | | |
| Male | 1440 (41.7) | 2608 (47.7) |
| Female | 2005 (58.1) | 2829 (51.8) |
| No response | 7 (0.2) | 25 (0.5) |
| Frailty status | | |
| Robust | 640 (19.0) | 750 (14.1) |
| Prefrailty | 1977 (58.7) | 3095 (58.3) |
| Frailty | 753 (22.3) | 1460 (27.5) |
| Subjective cognitive decline | | |
| No | 2367 (69.4) | 3529 (65.3) |
| Yes | 1042 (30.6) | 1878 (34.7) |
| Engagement in dementia prevention activities | | |
| Yes (doing) | 973 (28.6) | 931 (17.1) |
| No (not doing) | 2429 (71.4) | 4503 (82.9) |
| Internet environment at home | | |
| Yes | 2179 (63.6) | 2561 (47.3) |
| No | 1109 (32.4) | 2450 (45.2) |
| I don’t know | 139 (4.1) | 408 (7.5) |
| Experience using web conferencing systems | | |
| Yes | 494 (15.6) | 434 (8.5) |
| No | 2681 (84.4) | 4655 (91.5) |
| Experience using LINE | | |
| Yes | 2128 (64.6) | 2154 (41.9) |
| No | 1164 (35.4) | 2987 (58.1) |
| Experience using YouTube | | |
| Yes | 1219 (37.5) | 1166 (22.6) |
| No | 2033 (62.5) | 3997 (77.4) |

Data are presented as number (%).

Table S5: Responses to the question "If the program was distributed online, would you like to engage in it at home?" according to participant characteristics

| | If the program was distributed online, would you like to engage in it at home? | |
|---|---|---|
| | Yes | No |
| Age | | |
| ≤64 years | 155 (4.5) | 205 (3.7) |
| 65–74 years | 1274 (37.2) | 1685 (30.8) |
| 75–84 years | 1609 (47.0) | 2529 (46.2) |
| ≥85 years | 389 (11.4) | 1057 (19.3) |
| Sex | | |
| Male | 1534 (44.5) | 2532 (45.9) |
| Female | 1905 (55.3) | 2963 (53.7) |
| No response | 7 (0.2) | 25 (0.5) |
| Frailty status | | |
| Robust | 633 (18.8) | 757 (14.1) |
| Prefrailty | 1966 (58.4) | 3132 (58.5) |
| Frailty | 767 (22.8) | 1469 (27.4) |
| Subjective cognitive decline | | |
| No | 2334 (68.5) | 3577 (65.5) |
| Yes | 1073 (31.5) | 1884 (34.5) |
| Engagement in dementia prevention activities | | |
| Yes (doing) | 942 (27.6) | 987 (18.0) |
| No (not doing) | 2465 (72.4) | 4494 (82.0) |
| Internet environment at home | | |
| Yes | 2341 (68.4) | 2432 (44.4) |
| No | 974 (28.4) | 2607 (47.6) |
| I don't know | 110 (3.2) | 434 (7.9) |
| Experience using web conferencing systems | | |
| Yes | 516 (16.2) | 417 (8.2) |
| No | 2673 (83.8) | 4696 (91.8) |
| Experience using LINE | | |
| Yes | 2253 (68.0) | 2061 (39.9) |
| No | 1059 (32.0) | 3108 (60.1) |
| Experience using YouTube | | |
| Yes | 1363 (41.5) | 1036 (20.0) |
| No | 1918 (58.5) | 4152 (80.0) |

Data are presented as number (%).

Table S6: Responses to the question “If materials describing how to perform the program were provided, would you like to engage in it regularly at a club gathering?” according to participant characteristics

| | If materials describing how to perform the program were provided, would you like to engage in it regularly at the club gathering? | |
|---|---|---|
| | Yes | No |
| Age | | |
| ≤64 years | 137 (3.1) | 219 (4.9) |
| 65–74 years | 1419 (32.6) | 1507 (33.4) |
| 75–84 years | 2153 (49.5) | 1977 (43.8) |
| ≥85 years | 642 (14.8) | 812 (18.0) |
| Sex | | |
| Male | 1801 (41.1) | 2251 (49.5) |
| Female | 2570 (58.7) | 2274 (50.0) |
| No response | 10 (0.2) | 20 (0.4) |
| Frailty status | | |
| Robust | 775 (18.1) | 602 (13.6) |
| Prefrailty | 2492 (58.4) | 2594 (58.7) |
| Frailty | 1003 (23.5) | 1225 (27.7) |
| Subjective cognitive decline | | |
| No | 2989 (69.1) | 2913 (64.6) |
| Yes | 1336 (30.9) | 1594 (35.4) |
| Engagement in dementia prevention activities | | |
| Yes (doing) | 1205 (27.8) | 716 (15.8) |
| No (not doing) | 3122 (72.2) | 3806 (84.2) |
| Internet environment at home | | |
| Yes | 2506 (57.7) | 2224 (49.3) |
| No | 1626 (37.4) | 1952 (43.3) |
| I don’t know | 213 (4.9) | 335 (7.4) |
| Experience using web conferencing systems | | |
| Yes | 505 (12.5) | 419 (9.9) |
| No | 3532 (87.5) | 3808 (90.1) |
| Experience using LINE | | |
| Yes | 2370 (57.1) | 1892 (44.1) |
| No | 1777 (42.9) | 2397 (55.9) |
| Experience using YouTube | | |
| Yes | 1290 (31.3) | 1084 (25.3) |
| No | 2837 (68.7) | 3209 (74.7) |

Data are presented as number (%).

Table S7: Responses to the question "If materials describing how to perform the program were provided, would you like to engage in a program of about 10 min at each club gathering?" according to participant characteristics

| | If materials describing how to perform the program were provided, would you like to engage in a program of about 10 min at each club gathering? | |
|---|---|---|
| | Yes | No |
| Age | | |
| ≤64 years | 157 (3.3) | 199 (4.9) |
| 65–74 years | 1545 (32.4) | 1392 (33.9) |
| 75–84 years | 2345 (49.2) | 1794 (43.7) |
| ≥85 years | 720 (15.1) | 717 (17.5) |
| Sex | | |
| Male | 1940 (40.4) | 2091 (50.7) |
| Female | 2848 (59.3) | 2018 (48.9) |
| No response | 11 (0.2) | 19 (0.5) |
| Frailty status | | |
| Robust | 823 (17.6) | 563 (14.0) |
| Prefrailty | 2731 (58.5) | 2352 (58.5) |
| Frailty | 1115 (23.9) | 1106 (27.5) |
| Subjective cognitive decline | | |
| No | 3258 (68.8) | 2651 (64.8) |
| Yes | 1479 (31.2) | 1441 (35.2) |
| Engagement in dementia prevention activities | | |
| Yes (doing) | 1298 (27.4) | 623 (15.1) |
| No (not doing) | 3433 (72.6) | 3492 (84.9) |
| Internet environment at home | | |
| Yes | 2718 (57.2) | 2014 (49.1) |
| No | 1806 (38.0) | 1768 (43.1) |
| I don't know | 228 (4.8) | 317 (7.7) |
| Experience using web conferencing systems | | |
| Yes | 539 (12.2) | 381 (10.0) |
| No | 3889 (87.8) | 3443 (90.0) |
| Experience using LINE | | |
| Yes | 2560 (56.5) | 1718 (44.1) |
| No | 1974 (43.5) | 2175 (55.9) |
| Experience using YouTube | | |
| Yes | 1386 (30.7) | 998 (25.6) |
| No | 3123 (69.3) | 2900 (74.4) |

Data are presented as number (%).

Table S8: Association between results of the questionnaire on the dementia prevention program and participant characteristics (univariate logistic regression analysis)

| | Was aware of the Tottori Method Dementia Prevention Program | | Would like to try it at a club gathering if an instructor was dispatched [a] | | Would like to engage in the program at a club gathering if distributed online | | Would like to engage in the program at home if distributed online | | Would like to engage in the program regularly at a club gathering if materials were provided | | Would like to engage in about 10 min of the program at a club gathering if materials were provided | |
|---|---|---|---|---|---|---|---|---|---|---|---|---|
| | OR (95%CI) | P value | OR (95%CI) | P value | OR (95%CI) | P value | OR (95%CI) | P value | OR (95%CI) | P value | OR (95%CI) | P value |
| Age (years) | | | | | | | | | | | | |
| ≤64 | ref | | ref | | ref | | ref | | ref | | ref | |
| 65–74 | 1.08 (0.76–1.54) | 0.670 | 1.42 (1.14–1.78) | 0.002 | 1.15 (0.92–1.45) | 0.216 | 1.00 (0.80–1.25) | 1.000 | 1.51 (1.20–1.89) | <0.001 | 1.41 (1.13–1.76) | 0.003 |
| 75–84 | 1.32 (0.93–1.87) | 0.116 | 1.80 (1.45–2.25) | <0.001 | 1.10 (0.88–1.38) | 0.389 | 0.84 (0.68–1.05) | 0.120 | 1.74 (1.39–2.17) | <0.001 | 1.66 (1.33–2.06) | <0.001 |
| ≥85 years | 1.09 (0.75–1.58) | 0.645 | 1.40 (1.10–1.77) | 0.005 | 0.83 (0.57–0.92) | 0.009 | 0.49 (0.38–0.62) | <0.001 | 1.26 (1.00–1.60) | 0.053 | 1.27 (1.01–1.61) | 0.043 |
| Sex | | | | | | | | | | | | |
| Male | ref | | ref | | ref | | ref | | ref | | ref | |
| Female | 1.54 (1.36–1.76) | <0.001 | 1.57 (1.44–1.71) | <0.001 | 1.28 (1.18–1.40) | <0.001 | 1.06 (0.97–1.16) | 0.174 | 1.41 (1.30–1.54) | <0.001 | 1.52 (1.40–1.65) | <0.001 |
| No response | 1.27 (0.44–3.63) | 0.657 | 0.27 (0.11–0.68) | 0.005 | 0.51 (0.22–1.18) | 0.113 | 0.46 (0.20–1.07) | 0.072 | 0.63 (0.29–1.34) | 0.226 | 0.62 (0.30–1.31) | 0.215 |
| Frailty status | | | | | | | | | | | | |
| Robust | ref | | ref | | ref | | ref | | ref | | ref | |
| Prefrailty | 0.69 (0.58–0.81) | <0.001 | 0.85 (0.75–0.96) | 0.008 | 0.75 (0.66–0.84) | <0.001 | 0.75 (0.67–0.85) | <0.001 | 0.75 (0.66–0.84) | <0.001 | 0.79 (0.70–0.90) | <0.001 |
| Frailty | 0.65 (0.54–0.79) | <0.001 | 0.69 (0.60–0.79) | <0.001 | 0.60 (0.53–0.69) | <0.001 | 0.62 (0.54–0.72) | <0.001 | 0.64 (0.56–0.73) | <0.001 | 0.69 (0.60–0.79) | <0.001 |
| Subjective cognitive decline | | | | | | | | | | | | |
| No | ref | | ref | | ref | | ref | | ref | | ref | |
| Yes | 0.78 (0.68–0.89) | <0.001 | 0.92 (0.84–1.01) | 0.075 | 0.83 (0.76–0.91) | <0.001 | 0.87 (0.80–0.96) | 0.004 | 0.82 (0.75–0.89) | <0.001 | 0.84 (0.76–0.91) | <0.001 |
| Engagement in dementia prevention activities | | | | | | | | | | | | |
| Yes (doing) | ref | | ref | | ref | | ref | | ref | | ref | |
| No (not doing) | 0.30 (0.26–0.34) | <0.001 | 0.42 (0.37–0.47) | <0.001 | 0.52 (0.47–0.57) | <0.001 | 0.58 (0.52–0.64) | <0.001 | 0.49 (0.44–0.54) | <0.001 | 0.47 (0.42–0.53) | <0.001 |

Respondents with missing data in their survey responses were excluded from the analyses that utilized items with missing values.

[a] Analysis excluding participants who answered "I have already tried it."

Abbreviations: OR, odds ratio; CI, confidence interval.

Table S9: Association between results of the questionnaire on the dementia prevention program and home internet environment and the use of online tools (univariate logistic regression analysis)

| | Was aware of the Tottori Method Dementia Prevention Program | | Would like to engage in the program at a club gathering if distributed online | | Would like to engage in the program at home if distributed online | |
|---|---|---|---|---|---|---|
| | OR (95%CI) | P value | OR (95%CI) | P value | OR (95%CI) | P value |
| Internet environment at home | | | | | | |
| Yes | ref | | ref | | ref | |
| No | 0.71 (0.62–0.81) | <0.001 | 0.53 (0.49–0.58) | <0.001 | 0.39 (0.35–0.43) | <0.001 |
| I don't know | 0.47 (0.34–0.65) | <0.001 | 0.40 (0.33–0.49) | <0.001 | 0.26 (0.21–0.33) | <0.001 |
| Experience using web conferencing systems | | | | | | |
| Yes | ref | | ref | | ref | |
| No | 0.55 (0.46–0.66) | <0.001 | 0.51 (0.44–0.58) | <0.001 | 0.46 (0.40–0.53) | <0.001 |
| Experience using LINE | | | | | | |
| Yes | ref | | ref | | ref | |
| No | 0.63 (0.55–0.72) | <0.001 | 0.39 (0.36–0.43) | <0.001 | 0.31 (0.28–0.34) | <0.001 |
| Experience using YouTube | | | | | | |
| Yes | ref | | ref | | ref | |
| No | 0.72 (0.63–0.82) | <0.001 | 0.49 (0.44–0.54) | <0.001 | 0.35 (0.32–0.39) | <0.001 |

Respondents with missing data in their survey responses were excluded from the analyses that utilized items with missing values.
Abbreviations: OR, odds ratio; CI, confidence interval.

Table S10: Association between results of the questionnaire on the dementia prevention program and home internet environment and the use of online tools (multiple logistic regression analysis)

| | Was aware of the Tottori Method Dementia Prevention Program | | Would like to engage in the program at a club gathering if distributed online | | Would like to engage in the program at home if distributed online | |
|---|---|---|---|---|---|---|
| | aOR (95%CI) [a] | P value | aOR (95%CI) [a] | P value | aOR (95%CI) [a] | P value |
| Internet environment at home | | | | | | |
| Yes | ref | | ref | | ref | |
| No | 0.80 (0.67–0.95) | 0.011 | 0.79 (0.70–0.89) | <0.001 | 0.64 (0.57–0.73) | <0.001 |
| I don’t know | 0.48 (0.32–0.73) | <0.001 | 0.59 (0.46–0.76) | <0.001 | 0.48 (0.37–0.63) | <0.001 |
| Experience using web conferencing systems | | | | | | |
| Yes | ref | | ref | | ref | |
| No | 0.59 (0.47–0.73) | <0.001 | 0.77 (0.66–0.91) | 0.002 | 0.90 (0.76–1.05) | 0.182 |
| Experience using LINE | | | | | | |
| Yes | ref | | ref | | ref | |
| No | 0.77 (0.65–0.93) | 0.006 | 0.54 (0.48–0.61) | <0.001 | 0.48 (0.43–0.55) | <0.001 |
| Experience using YouTube | | | | | | |
| Yes | ref | | ref | | ref | |
| No | 0.92 (0.76–1.11) | 0.362 | 0.73 (0.64–0.83) | <0.001 | 0.61 (0.53–0.69) | <0.001 |

Respondents with missing data in their survey responses were excluded from the analyses that utilized items with missing values.

[a] Analysis in which one of the following variables was used as the explanatory variable and the remaining variables adjusted for as covariates: age, sex, frailty assessment, subjective cognitive decline assessment, engagement in dementia prevention activities, internet environment at home, and experience using web conferencing systems, LINE, or YouTube.

Abbreviations: aOR, adjusted odds ratio; CI, confidence interval.

Appendix: STROBE Statement—Checklist of items that should be included in reports of ***cross-sectional studies***

| | Item No | Recommendation | Section |
|---|---|---|---|
| **Title and abstract** | 1 | (*a*) Indicate the study's design with a commonly used term in the title or the abstract | Title |
| | | (*b*) Provide in the abstract an informative and balanced summary of what was done and what was found | Abstract |
| **Introduction** | | | |
| Background/rationale | 2 | Explain the scientific background and rationale for the investigation being reported | Introduction |
| Objectives | 3 | State specific objectives, including any prespecified hypotheses | Introduction |
| **Methods** | | | |
| Study design | 4 | Present key elements of study design early in the paper | Methods, Study design |
| Setting | 5 | Describe the setting, locations, and relevant dates, including periods of recruitment, exposure, follow-up, and data collection | Methods, Participants and data collection |
| Participants | 6 | (*a*) Give the eligibility criteria, and the sources and methods of selection of participants | Methods, Participants and data collection |
| Variables | 7 | Clearly define all outcomes, exposures, predictors, potential confounders, and effect modifiers. Give diagnostic criteria, if applicable | Methods, Questionnaire survey |
| Data sources/ measurement | 8* | For each variable of interest, give sources of data and details of methods of assessment (measurement). Describe comparability of assessment methods if there is more than one group | Methods: Questionnaire survey |
| Bias | 9 | Describe any efforts to address potential sources of bias | N/A |
| Study size | 10 | Explain how the study size was arrived at | Methods: Participants and data collection |
| Quantitative variables | 11 | Explain how quantitative variables were handled in the analyses. If applicable, describe which groupings were chosen and why | N/A |
| Statistical methods | 12 | (*a*) Describe all statistical methods, including those used to control for confounding | Methods: Statistical analysis |
| | | (*b*) Describe any methods used to examine subgroups and interactions | N/A |
| | | (*c*) Explain how missing data were addressed | Methods: Participants and data collection |
| | | (*d*) If applicable, describe analytical methods taking account of sampling strategy | N/A |
| | | (*e*) Describe any sensitivity analyses | Methods: Statistical analysis |

| | | | |
|---|---|---|---|
| **Results** | | | |
| Participants | 13* | (a) Report numbers of individuals at each stage of study—eg numbers potentially eligible, examined for eligibility, confirmed eligible, included in the study, completing follow-up, and analysed | Methods: Participants and data collection |
| | | (b) Give reasons for non-participation at each stage | N/A |
| | | (c) Consider use of a flow diagram | N/A |
| Descriptive data | 14* | (a) Give characteristics of study participants (eg demographic, clinical, social) and information on exposures and potential confounders | Table 1 |
| | | (b) Indicate number of participants with missing data for each variable of interest | Table 1 |
| Outcome data | 15* | Report numbers of outcome events or summary measures | N/A |
| Main results | 16 | (*a*) Give unadjusted estimates and, if applicable, confounder-adjusted estimates and their precision (eg, 95% confidence interval). Make clear which confounders were adjusted for and why they were included | Table 2 and Supplementary Table S8 |
| | | (*b*) Report category boundaries when continuous variables were categorized | N/A |
| | | (*c*) If relevant, consider translating estimates of relative risk into absolute risk for a meaningful time period | N/A |
| Other analyses | 17 | Report other analyses done—eg analyses of subgroups and interactions, and sensitivity analyses | N/A |
| **Discussion** | | | |
| Key results | 18 | Summarise key results with reference to study objectives | Discussion: paragraph 1 |
| Limitations | 19 | Discuss limitations of the study, taking into account sources of potential bias or imprecision. Discuss both direction and magnitude of any potential bias | Discussion: paragraph 6 |
| Interpretation | 20 | Give a cautious overall interpretation of results considering objectives, limitations, multiplicity of analyses, results from similar studies, and other relevant evidence | Discussion |
| Generalisability | 21 | Discuss the generalisability (external validity) of the study results | Discussion: paragraph 6 |
| **Other information** | | | |
| Funding | 22 | Give the source of funding and the role of the funders for the present study and, if applicable, for the original study on which the present article is based | Important disclosures and Sponsor's role |

*Give information separately for exposed and unexposed groups.

**Note:** An Explanation and Elaboration article discusses each checklist item and gives methodological background and published examples of transparent reporting. The STROBE checklist is best used in conjunction with this article (freely available on the Web sites of PLoS Medicine at http://www.plosmedicine.org/, Annals of

Internal Medicine at http://www.annals.org/, and Epidemiology at http://www.epidem.com/). Information on the STROBE Initiative is available at www.strobe-statement.org.
